# Does cardiorespiratory fitness mediate or moderate the association between mid-life physical activity and cognitive function? Findings from the 1958 British birth cohort study

**DOI:** 10.1101/2023.11.15.23298607

**Authors:** Tom Norris, John J Mitchell, Joanna M Blodgett, Mark Snehal M Pinto Pereira

## Abstract

**Background:** Physical activity (PA) is associated with a lower risk of cognitive decline and all-cause dementia in later life. Pathways underpinning this association are unclear but may involve either mediation and/or moderation by cardiorespiratory fitness (CRF).

**Methods:** Data on participation in PA (exposure) at 42y, non-exercise testing CRF (NETCRF, mediator/moderator) at 45y and overall cognitive function (outcome) at 50y were obtained from 9,385 participants in the 1958 British birth cohort study. We used a four-way decomposition approach to examine the relative contributions of mediation and moderation by NETCRF on the association between PA at 42y and overall cognitive function at 50y.

**Results:** In males, the estimated overall effect of 42y PA ≥*once per week (vs. <once per week*) was a 0.08 (95% confidence interval: 0.04,0.13) higher overall cognitive function z-score at 50y. The estimated controlled direct effect was similar (0.08 (0.03, 0.12)). Thus, the proportion of the estimated effect via mediation or moderation by NETCRF was small (∼3%), with confidence intervals straddling the null. In females, there was no estimated overall effect of PA on overall cognitive function.

**Conclusion:** We present the first evidence from a four-way decomposition analysis of the potential contribution that CRF plays in the relationship between mid-life PA and subsequent cognitive function. Our lack of evidence in support of CRF mediating or moderating the PA―cognitive function association suggests that other pathways underpin this association.

## Background

Cognitive impairment, especially in later-life, is a public health concern because it raises the risk of developing various dementias(1). Dementia is a major cause of disability among older people, accounting for 11.9% of years lived with disability due to non-communicable diseases(2). As a result of population ageing, identifying strategies to alleviate cognitive impairment (and therefore dementia) will become even more critical(2). In the United Kingdom (UK), there are no effective treatments to reverse or delay dementia progression(3). Therefore, identifying modifiable causal factors which can be intervened upon to delay age-related declines in cognitive function is essential.

Physical activity (PA) is one such modifiable factor. Higher levels of PA are associated with lower risk of all-cause dementia(4, 5), cognitive decline(6) and better later-life cognition(7). However, pathways underpinning the PA―cognition association are unclear. While cardiovascular(8) and mental(5) health may be involved, an alternative pathway may include cardiorespiratory fitness (CRF). CRF is a marker of the capacity of the cardiovascular system to transport oxygen and the ability of muscle tissue to utilise it(9). PA, particularly aerobic activity, has beneficial effects on CRF(10) and evidence suggests that higher CRF is associated with greater subsequent cognitive function(11). Thus, CRF may have a mediating role in the PA―cognition association. Additionally, CRF may also moderate the PA―cognition association, such that associations differ depending on fitness levels as higher fitness would enable participation in greater volumes/intensity of PA. However, evidence is mixed and may be specific to certain cognitive domains. For example, some studies suggest that the effect of PA on memory is stronger in fitter individuals(12), whilst others find the positive effect of PA on processing speed is stronger in less-fit individuals(13).

It is plausible that CRF exerts both mediating and moderating effects on the PA―cognition association. Conventional analyses that ignore the potential mediation and moderation of PA by CRF on cognitive outcomes can therefore be biased(14). Thus, the ability to incorporate both these effects into analyses is critical for obtaining a more accurate insight into underlying mechanisms through which PA influences cognition. To our knowledge, no study has investigated the mechanisms underlying the PA―cognition association considering potential mediation and moderation by CRF simultaneously. Therefore, we attempt to address this critical knowledge gap using a four-way decomposition approach(15) in an age-homogenous general population sample born in 1958 and followed-up throughout their lives. Our objective was to formally examine the relative contribution of mediation and moderation by CRF on the PA―cognition association.

## Methods

Data come from the 1958 British birth cohort(16), which enrolled 17,638 participants at birth during a single week in March 1958 in Great Britain and subsequently added a further 920 immigrants born in the same week. In mid-adulthood (45 years (y)), participants remain broadly representative of the original study sample(17). At 42y (n=11,419) and 50y (n=9,790) participants completed face-face interviewer-administered questionnaires in their homes. At 45y (n=9,377) they were visited by a nurse who collected biomedical, physical and sociodemographic information.

Our analytic sample (N=9,385) includes participants with valid measures on all cognitive tests undertaken at 50y (see Supplementary figure S1). Ethical approval was given by the London multi-centre research ethics committee for the age 42y (reference not available) and 50y (08/H0718/29) sweeps, whilst the South-East multi-centre research ethics committee (01/1/44) provided ethical approval for the age 45y sweep. Written informed consent was obtained from participants at all ages. Full details are available elsewhere(16, 18). Data were accessed for research purposes on 27/8/21. Authors had no access to information that could have identified individual participants during or after data collection.

### *Exposure*: Physical activity frequency (42y)

At 42y, participants were asked whether, and how often they participated in a range of leisure-time physical activities (e.g., competitive sports of any kind, keep-fit classes, running; further details in supplementary text S1). We derived a dichotomous variable representing whether people participated in leisure-time physical activities ‘*less than once per week’* (0) or *‘at least once per week’* (1).

### *Mediator*: Non-exercise testing cardiorespiratory fitness (NETCRF, 45y)

At 45y (range 44y-46y), NETCRF was predicted using information on sex, age, body mass index (BMI), resting heart rate (RHR), and self-reported PA. The prediction equation used has strong validity against exercise testing-estimated fitness(19). Age was recorded in whole years. Weight and height were measured using standard protocols(20); BMI (kg/m^2^) was calculated. After a few minutes rest, three measurements of RHR were obtained (taken one minute apart) using an automated device (Omron 907 blood pressure monitor, Omron Healthcare, UK); mean RHR was calculated. A modified version of the European Prospective Investigation into Cancer and Nutrition Physical Activity Questionnaire (EPIC-PAQ) was used to assess leisure-time PA(21).

NETCRF was calculated as(22):

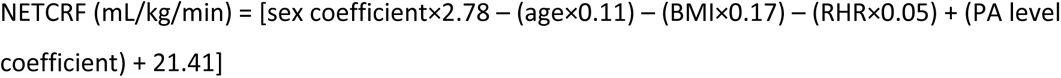

Where, the sex coefficient was 1 for males and 0 for females. The PA coefficients were based on adherence to contemporaneous guidelines(22) assessed using EPIC-PAQ: 0.0 for inactive during leisure-time; 0.29 for active, but not meeting guidelines; and, 1.21 for meeting guidelines (i.e., at least 150minutes/week moderate-intensity or 75minutes/week vigorous-intensity PA). NETCRF estimates were converted into maximal aerobic capacity metabolic equivalent (MET) values (1 MET corresponds to an oxygen consumption of 3.5 mL/kg/min (based on a 40y male, weighing 70kg)(22)).

### *Outcome*: Overall cognitive function (50y)

‘Overall cognitive function’ was calculated as the average score of four cognitive function tasks completed at 50y(23). The four tasks, completed in the following order, were: i)immediate verbal memory; ii)verbal fluency; iii)visual processing speed; and iv)delayed verbal memory. These tests have been routinely used as measures of cognition in large-scale epidemiological studies(24) and are predictive of incident dementia(25). For immediate verbal memory, participants were played an audio recording of 10 words and were given two minutes to orally recall them. Verbal fluency was assessed via an animal naming test, in which respondents were given one minute to name as many animals as they could think of. Visual processing speed was assessed using a dual-letter cancellation test, in which participants were presented with blocks of letters and were asked to read through the blocks from left to right, crossing out ‘Ws’ and ‘Ps’ as fast as they could. Search speed was calculated by summing the total number of letters scanned, including both target and non-target letters. Delayed recall was measured by asking participants to recall as many words as they could from the original list presented to them during the first word-recall task, with a two-minute cut-off. The four measures were standardised (z-score, separately for each sex) and the average (overall cognitive function) was calculated.

#### Potential confounders

Potential baseline and intermediate confounders of the PA-CRF-cognitive function association were identified a-priori and included in the directed acyclic graph (Supplementary figure S2). They included: Social class at birth, childhood cognitive function (11y), educational attainment (33y), smoking status (42y), alcohol consumption (42y), BMI (45y), PA level (45y), sleep problems (45y) and self-rated health over the previous 12 months (45y); details in Supplementary materials.

#### Statistical analysis

Due to observed sex differences in the PA―cognition association(26), and the CRF distribution(27), we chose a-priori to perform sex-stratified analyses.

Regression models were used to explore the relationship between i) PA at 42y and cognitive function at 50y; ii) PA at 42y and NETCRF at 45y; and iii) NETCRF at 45y and cognitive function at 50y. Models were first unadjusted and then adjusted for baseline confounders (the NETCRF―cognition association was additionally adjusted for PA at 42y). To address missingness in covariate data (see Table 1), we used multiple imputation by chained equations under a missing-at-random assumption; the number of imputations (n=30) required to achieve convergence of parameter estimates was determined as 100×fraction missing information(28). All variables described above (i.e., 42y PA, 45y NETCRF, 50y cognitive function and potential confounders) were included in the imputation model, as well as childhood internalizing and externalizing behaviours and cognitive ability which have been shown to predict missingness in follow-up(17).

**Table 1.**
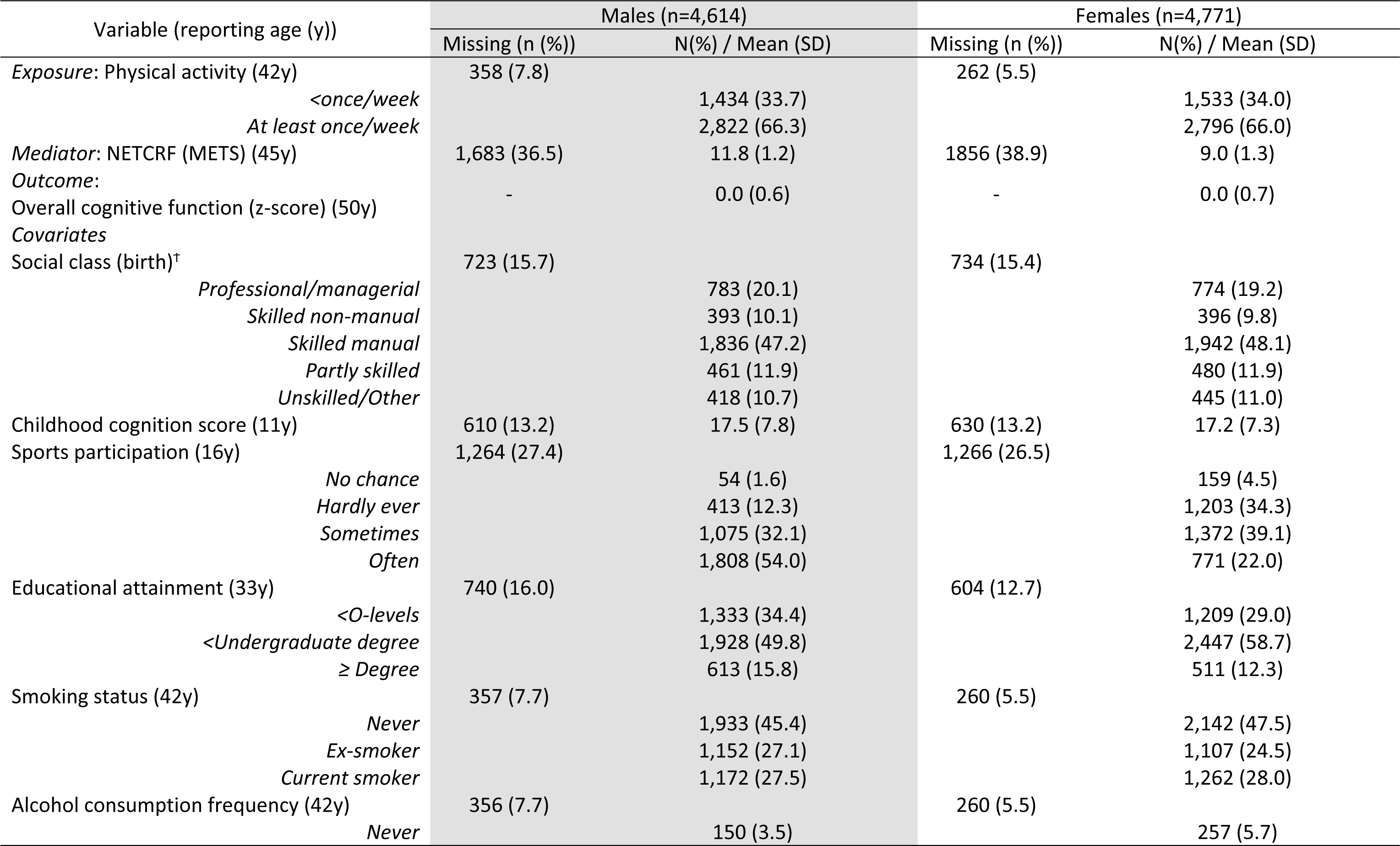
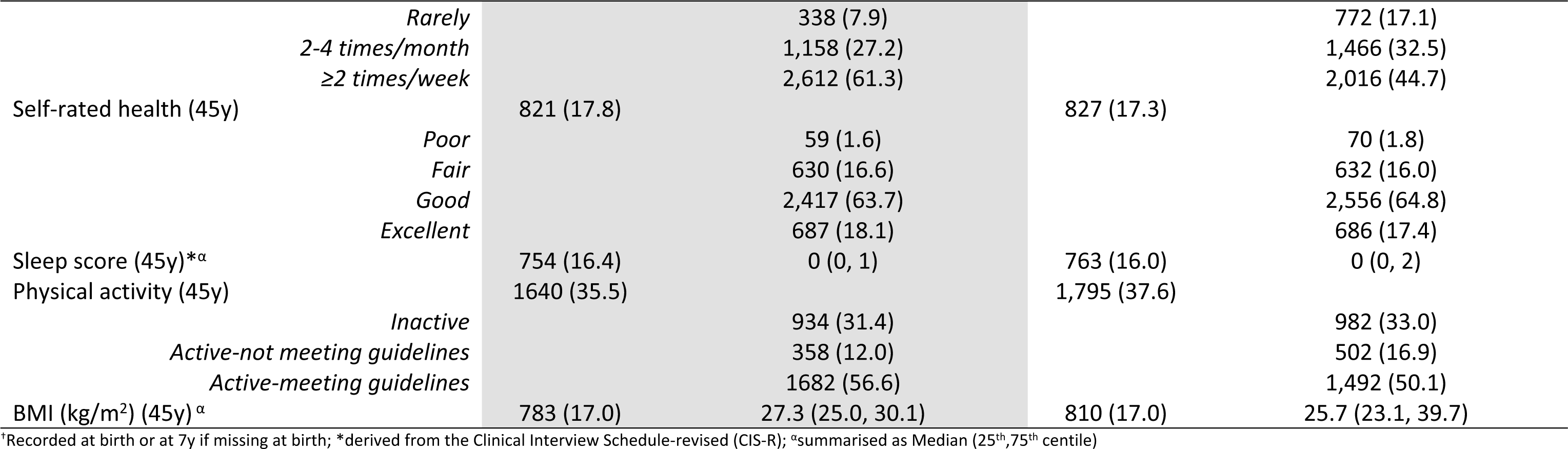
Sample characteristics (n=9,385)

#### 4-way decomposition analysis

To estimate the relative contribution of the potential pathways linking mid-adult PA to subsequent cognitive function, we used a four-way decomposition analysis. This methodology allows us to assess the extent to which the estimated overall effect (_e_OE) of PA on cognitive function is explained by alternative pathways involving (or not involving) NETCRF. Specifically, the method decomposes the estimated overall PA―cognitive function effect into 4 sub-components: an estimated controlled direct effect (_e_CDE); an estimated randomised analogue of the pure indirect effect (_e_rPNIE); an estimated randomised analogue of the reference interaction (_e_rINT_REF_) and an estimated randomised analogue of the mediated interaction (_e_rINT_MED_). Briefly, _e_CDE is the portion of the _e_OE of PA on cognitive function due to pathways that do not involve NETCRF (neither mediation nor interaction). _e_rPNIE is the estimated effect only due to the mediation pathway that does not involve interaction. _e_rINT_REF_ is the estimated effect of PA on cognitive function which is due solely to the interaction between PA and NETCRF (i.e., effect of PA varies by fitness level). _e_rINT_MED_ is the estimated effect of PA on cognitive function due to both the interaction between PA and NETCRF and the mediating effect of NETCRF.

The four sub-components sum up to the _e_OE of PA on cognitive function. In addition to the four-way decomposition of the _e_OE described above, we also calculated randomised analogues for the proportions mediated by NETCRF, attributable to the PA-NETCRF interaction and the proportion eliminated (due to both mediation by, and interaction between NETCRF and PA). Notably, we account for intermediate confounders of the NETCRF―cognitive function association that are affected by PA (e.g., BMI), by estimating randomised analogue of effects. All effects were estimated on the difference scale, with 95% confidence intervals (CIs) calculated from bootstrapped standard errors (n=100). See Figure 1 and supplementary table S1 for further details.

**Figure 1.**
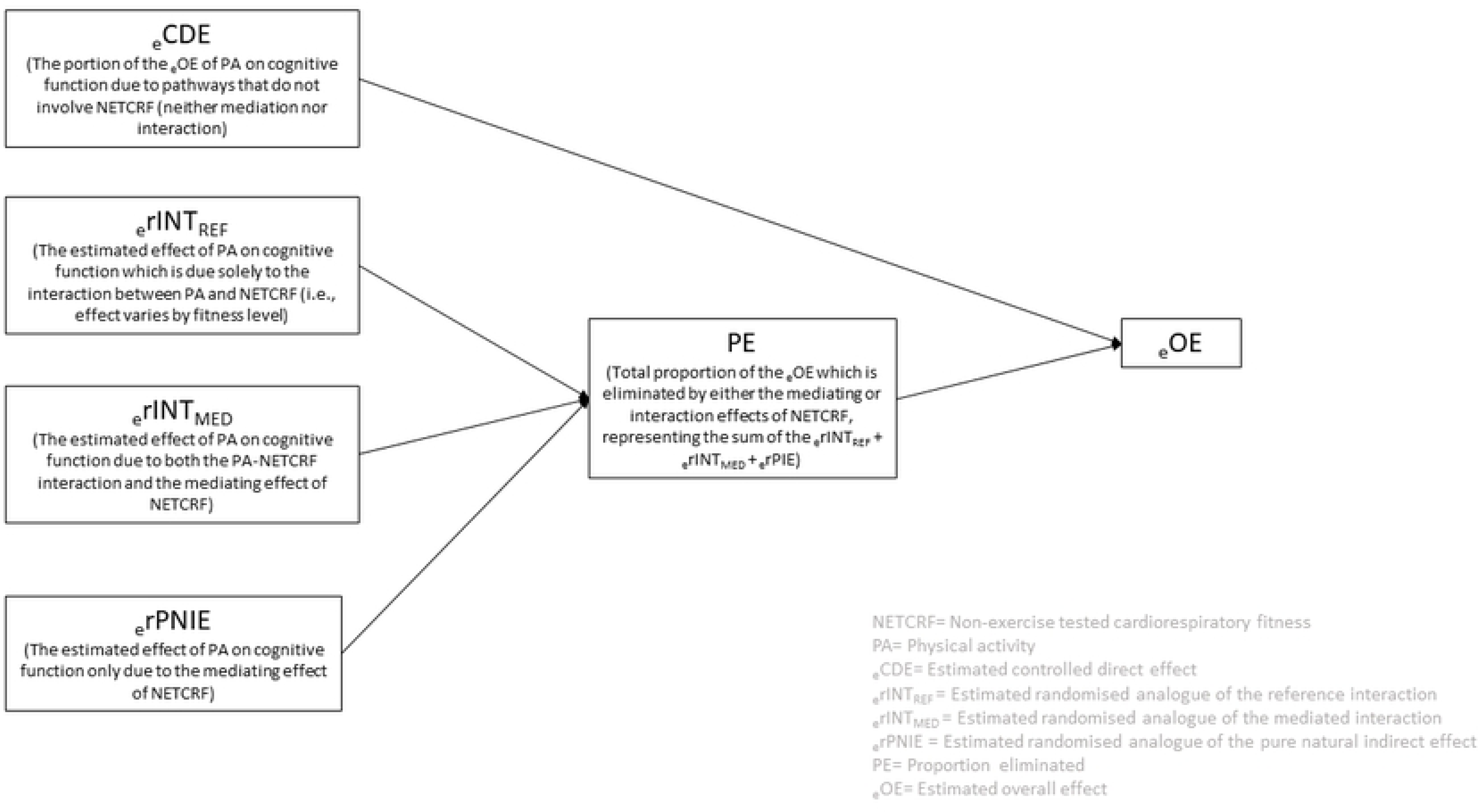
Four-way decomposition of the estimated effect of PA at 42y on cognitive function at 50y

#### Supplementary analyses

To identify the extent to which unmeasured confounding may influence estimates, we used the E-value approach of Vanderweele & Ding(29). An E-value is the minimum strength of association an unmeasured confounder would need to have with both PA and cognitive function, conditional on measured covariates, to fully explain away the PA–cognitive function association. Our estimated causal effects were on the difference scale; thus, they were transformed into risk ratios using the Vanderweele & Ding transfomation(29) before E-values were calucated. We report the (i) E-value of the estimate and (ii) E-value for the limit of the CI closest to the null.

To determine whether effects varied by cognitive function domains, we repeated the four-way decomposition analysis using each of the 4 individual cognitive function tasks as our outcome. Analyses were conducted in Stata MP v15.1 and R 4.3.0 (using the ‘*CMAverse’*(*30*) suite of functions).

## Results

### Sample characteristics

Approximately two thirds of males and females reported being physically active at least once/week at 42y (Table 1). On average, NETCRF at 45y was slightly higher in males (11.8 METS (standard deviation (SD): 1.2) vs 9.0 METS (SD:1.3)), whereas females performed slightly better on all four cognitive tasks at 50y (Supplementary table S2).

### Association between 42y PA, 45y NETCRF and 50y overall cognitive function

42y PA was associated with higher 45y NETCRF; e.g., after adjusting for baseline confounders, males doing PA at least once/week had 0.44METS (95% CI: 0.36,0.53) higher NETCRF than those engaging less than once/week (Table 2). 42y PA was also associated with 50y overall cognitive function; e.g., in males, after adjusting for baseline confounders, PA at least once/week was associated with a 0.08 (95% CI: 0.04, 0.11) higher overall cognitive function z-score. Finally, while a 1-unit higher 45y NETCRF in both sexes was associated with higher 50y overall cognitive function in unadjusted models, associations were fully attenuated after adjustment for baseline confounders and 42y PA.

**Table 2.**
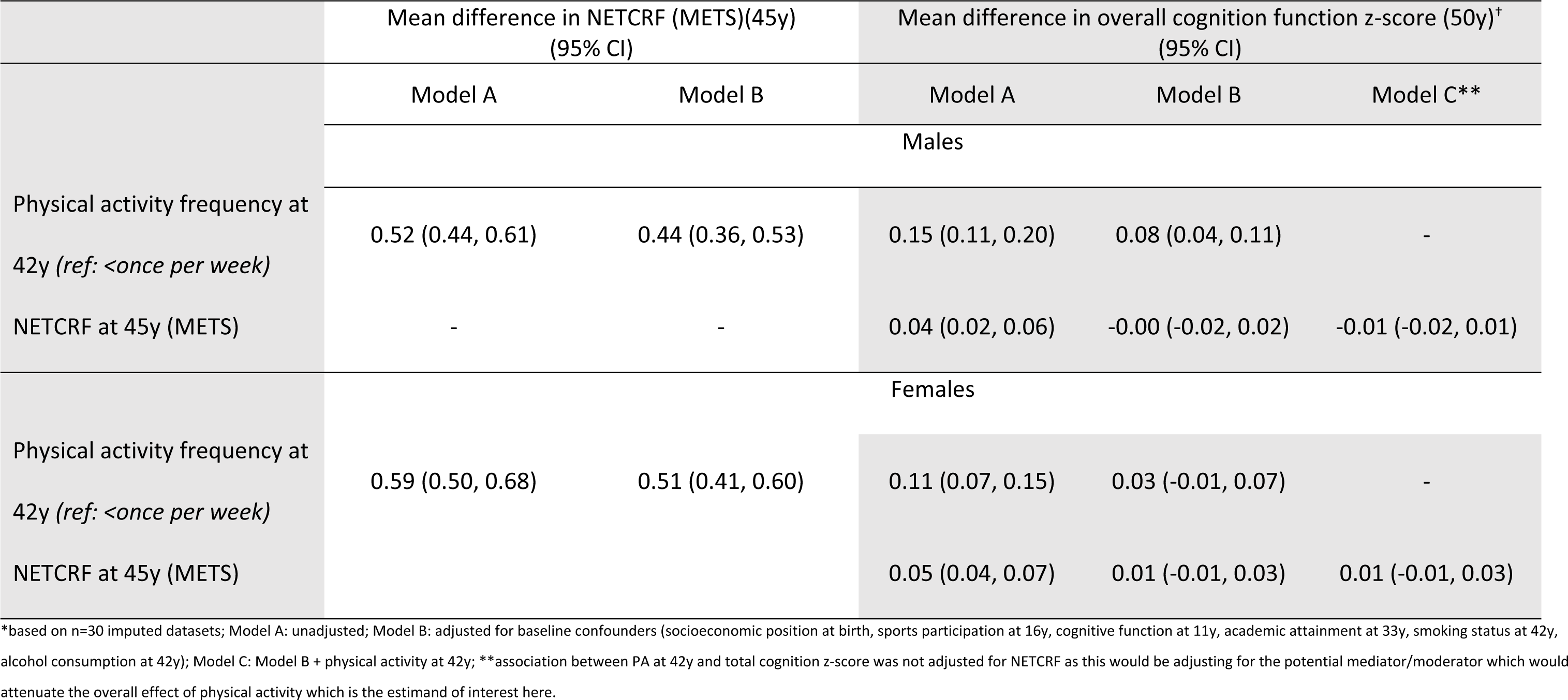
Associations between PA, NETCRF and overall cognitive function (N=9,385)*

### 4-way decomposition analysis

#### Males

There was a small positive _e_OE of 42y PA on 50y overall cognitive function (expressed as a difference in mean z-scores); i.e., the _e_OE of PA at least once/week at 42y (vs. less than once/week) was a 0.08 (95% CI: 0.04,0.13) higher 50y overall cognitive function z-score. The _e_CDE was 0.08 (0.03,0.12), indicating that the PA―overall cognitive function association was mainly due to pathways excluding NETCRF. Thus, the _e_rPNIE, _e_rINTREF and _e_rINTMED effects were all consistent with the null (Table 3).

**Table 3.** Estimated overall, controlled direct and randomised analogues of the pure indirect, reference interaction and mediated interaction effects* of PA at 42y on overall cognitive function at 50y (mediated/moderated by NETCRF at 45y)

|  | Difference in mean overall cognitive function z-score (95% CI) |  |
| --- | --- | --- |
|  | Physical activity (42y; ref: PA<once/week) |  |
|  | Males (n=4,614) | Females (n=4,771) |
| Overall effect (εOE) | 0.08 (0.04, 0.13) | 0.03 (-0.01, 0.07) |
| Controlled Direct Effect (εCDE) | 0.08 (0.03, 0.12) | 0.02 (-0.02, 0.07) |
| Randomized analogue of Pure Natural Indirect Effect (εrPNIE) | 0.01 (-0.01, 0.03) | 0.02 (-0.00, 0.04) |
| Randomized analogue of Reference Interaction (εrINTREF) | -0.00 (-0.00, 0.00) | -0.00 (-0.00, 0.00) |
| Randomized analogue of Mediated Interaction (εrINTMED) | -0.01 (-0.02, 0.01) | -0.01 (-0.02, 0.00) |
| Randomized analogue of overall proportion mediated (εrPM) | 0.05 (-0.11, 0.31) | 0.36 (-1.98, 2.68) |
| Randomized analogue of overall proportion attributable to interaction (εrINT) | -0.10 (-0.32, 0.12) | -0.27 (-1.71, 0.63) |
| Randomized analogue of overall proportion eliminated (εrPE) | 0.03 (-0.12, 0.31) | 0.33 (-1.75, 2.44) |

\* $\epsilon$ CDE is the portion of the $\epsilon$ OE of PA on cognitive function due to pathways that do not involve NETCRF (neither mediation nor interaction); $\epsilon$ rPNIE is the effect only due to the mediation pathway that does not involve interaction; $\epsilon$ rINTREF is the effect of PA on cognitive function which is due solely to the interaction between PA and NETCRF; $\epsilon$ rINTMED is the effect of PA on cognitive function due to the effect of NETCRF as both a mediating and moderating variable. The four sub-components sum up to the $\epsilon$ OE of PA on cognitive function: see methods and supplementary materials for details.

#### Females

The _e_OE of PA at 42y on overall cognitive function at 50y was small and confidence intervals straddled the null (0.03 (95% CI: −0.01, 0.07)). While there was some evidence of mediation by NETCRF, all four estimated decomposed effects were consistent with the null (Table 3).

### Supplementary analyses

The E-values for the causal estimates are presented in Supplementary Table S3. In males, the E-values suggest that the risk ratios for the _e_OE (RR=1.11) and _e_CDE (RR=1.11) could be fully explained by an unmeasured confounder that was associated with both PA at 42y and overall cognitive function at 50y by a risk ratio of approximately 1.47 and 1.46, respectively, independent of the measured confounders. The lower limit of the 95% CI could be moved to include the null by an unmeasured confounder that was associated with PA and overall cognitive function by a risk ratio of approximately 1.29 and 1.27, respectively. In females, smaller amounts of unmeasured confounding would be required to explain away the estimated effect of PA at 42y on overall cognitive function at 50y. Furthermore, because the confidence interval of the RR already straddled the null, unmeasured confounding was unneccessary to move the CI to the null.

Associations reported above for overall cognitive function in males, was driven mainly by associations between PA and immediate and delayed verbal memory (Supplementary Tables S4-S7). For females, as per the main analysis, there was no estimated effect of PA on any of the four cognitive function sub-domains.

## Discussion

Using data from a large general population sample followed from birth for over five decades, we present novel findings, obtained from an under-utilised four-way decomposition approach, regarding the potential mediating and/or moderating effects of CRF on the relationship between PA at 42y and overall cognitive function 8 years later. In males, we observed a small positive estimated overall effect of PA on subsequent overall cognitive function, which was almost exclusively driven by an estimated direct effect of PA, with no strong evidence in support of CRF either mediating or moderating the association. In females, once confounders were considered, there was no estimated overall effect of PA on overall cognitive function.

While there is a general consensus that engagement in PA is beneficial for cognition(31), evidence of the benefits of PA engagement in mid-life is mixed, with some studies citing a lack of evidence(31), whilst others report beneficial effects(7). In contrast to our findings, a recent study in 1,147 participants born in Britian in 1946, observed consistent positive associations between participation in lesisure time PA from 36y onwards, and cognitive function at 69y, though estimates were smallest and attenuated when looking only at PA engagement in early-to-mid-adulthood (36-43y)(32). As different PA and cognitive function measures, assessed at a later life-stage (69y vs 50y), were used, drawing comparisons between the studies is difficult. However, as was observed in our study, it may be that PA only results in small benefits to subsequent cognitive function and thus sustained PA over a longer time period may be required in order accumulate these small incremental benefits that translate into tangible improvements in cognitive function.

We found evidence that the PA―cognition relationship may be sex-specific, observing in males, a small positive estimated effect of PA at 42y and overall cognitive function 8 years later, but no effect in females. While we did observe an association between PA at 42y in females and subsequent cognitive function in unadjusted models, this association was attenuated to the null upon adjustment for baseline confounders, suggesting a potential important confounding pathway in females. This is in contrast to a recent report from the English Longitudinal Study of Ageing, which found evidence for a relationship between PA and cognitive decline in females, but not in males(33). Furthermore, evidence from randomised trials (which are less likely to be affected by confounding variables) in older participants (average age >70y) with a cognitive impairment diagnosis, also suggests that the relationship between PA and cognitive function is greater in females(26, 34). Several mechanisms have been proposed to explain possible sex-specific effects of PA engagment on later cognition, including differences in: neuroplasticity, brain-derived neurotrophic factor (BDNF) levels and physiological adaptations to exercise(26).

Our work is novel because, to our knowledge, no other study has examined simultaneously the mediating and moderating role of CRF on the relationship between mid-life PA and subsequent cognitive function. Our formal mediation analysis is in line with a 2015 Cochrane review(35), which found no evidence that PA-induced increases in CRF resulted in improved cognitive performance in healthy older adults without known cognitive impairment. More recently, studies in children have observed a mediating effect of fitness on the relationship between PA and executive function(36) and academic attainment(37). However, these studies, in children aged 9-13y, may not be generalisable to adults. Studies in adults report mixed findings, with some studies finding evidence of a mediating effect of fitness(38), whilst others have not(39). If positive effects of mid-life PA on cognitive function are not a result of intermediate increases in fitness, what may be the mode of action? Proposed mechanisms include a reduced likelihood of vascular diseases and improvements in cerebral perfusion(40–42), stimulation of growth factors (e.g., brain-derived neurotrophic factor(43)), and/or the downregulation of oxidative stress and inflammatory responses(44). Alternatively, PA may positively affect cognitive function, independent of the movement component of exercise, due to the cognitively stimulating aspects of PA, such as eye-hand coordination, visuospatial memory, self-motivation(45), planning and social interaction(46, 47).

In trying to explain the apparent lack of a *PA-CRF-cognitive outcome* mechanism of action, Young et al (2015) suggested that PA effects may operate in certain subgroups of the population only, such as those starting from a lower baseline of CRF, i.e., a moderating effect may exist between PA and CRF(35). A major novelty of our study, due to the use of the 4-way decomposition approach(15), is that we have been able to investigate the contribution that any PA-CRF interaction effect exerts on cognitive function. We observed no evidence of an interaction effect, such that in males, the positive effect of PA at 42y was consistent across the distributon of NETCRF at 45y. Previous studies have reported mixed findings regarding the interactive effect of fitness on the relationship between PA and cognitive outcomes, with some observing evidence of a greater benefit of PA in those with higher fitness(12), lower fitness(13), or no effect(48).

Our study has a number of strengths. For example, our use of a four-way decomposition analysis, has allowed us, for the first time, to more accurately reflect likely processes in the real world, thus avoiding potential biases inherent in simpler methods. We were able to account for intermediate confounders (e.g., BMI at 45y) which are themselves predicted by our exposure (i.e., PA at 42y). Importantly, our methodology allowed us to robustly examine mediation and moderation simultaneously. This was possible due to the cohort’s prospective design with multiple follow-up time points, enabling us to respect the temporal ordering of our exposure, mediator, outcome and confounding variables. Thus the possibility of reverse causation is reduced. Our outcome, overall cognitive function, was assessed at approximately the same age (50y) for all individuals, thus removing the known influence of age on cognitive function(49). Our overall cognitive function variable was based on the average of four cognitive function tasks which have demonstrated validity in the prediction of incident dementia(50) and as a supplementary analysis we examined each cognitive domain seperately. Nonetheless we acknowledge study limitations. For example, our approach relies on several assumptions including no unmeasured confounding of the PA-cognitive function, PA-NETCRF and NETCRF-cognitive function associations. While availability of detailed, prospectively collected covariate data enabled us to account for several important confounders, the possibility of residual confounding could not be ruled out. Therefore, we investigated the extent to which unmeasured confounding may be influencing our estimates, using E-values(29). Our binary exposure, PA at 42y, was based on self-report and may be subject to measurement error and bias and, due to data limiations, were unable to consider duration and intensity of PA. Our mediator, NETCRF at 45y, was predicted using an equation that has been well validated in adults(19). While it is an appropriate proxy for CRF in large scale studies, such as the 1958 birth cohort, we acknowledge that NETCRF is not equivalent to the gold-standard assessment of CRF via tests to exhaustion measuring oxygen uptake(10). Furthermore, our average NETCRF estimates indicate a relatively fit sample when compared to adults of a similar age in the general population(51). As in all longitudinal studies, loss to follow-up occurred and while participants in mid-adulthood were broadly representative of the original population, the most disadvantaged were least likely to remain(17). Finally, the 1958 birth cohort is predominantly of White British ethnicity (∼98% at 45y). For the above reasons, generalisability (e.g., to other ethnic groups, less fit middle-aged men and women) should be infered with caution.

We provide the first evidence obtained regarding the simultaneously mediating and moderating effect of CRF on the relationship between mid-life PA and subsequent cognitive function. We observed an estimated total and direct effect of PA at 42y on overall cognitive function at 50y in males only. In both sexes, there was no mediation or moderation of PA operating via fitness, suggesting that any positive effect of PA on cognitive function operates through other mechanisms which need to be explored.

## Acknowledgements

The authors are grateful to the Centre for Longitudinal Studies (CLS), UCL Institute of Education, for the use of the 1958 cohort data and to the UK Data Service for making them available. However, neither CLS nor the UK Data Service bear any responsibility for the analysis or interpretation of these data.

## Declarations

## Data Availability

The original data for the 1958 NCDS are available from the UK Data Service; applications for access to any data held by the UK Data Archive that forms part of the NCDS Biomedical Resource will require special license and should be submitted to.

